# Utility of a Clinical Scoring System for Point of Care Triaging in COVID-19 Pneumonia

**DOI:** 10.1101/2021.02.26.21252256

**Authors:** Andrew J Gangemi, Rohit Gupta, Gustavo Fernandez-Romero, Huaqing Zhao, Maulin Patel, Junad Chowdhury, Massa Zantah, Matthew Zheng, Osheen Abramian, Stephen Codella, Linda Vien, Eduardo Dominguez-Castillo, Timothy Buckey, Charles Earley, Jourdan Frankovich, Mali Jurkowski, Zachary Jurkowski, Jenny O’Brien, Nanzhou Guo, Paige Stanley, Brenton Halsey, Jasleen Kahlon, Navjot Kaur, Roman Prosniak, Maruti Kumaran, Chandra Dass, David Fleece, Michael R Jacobs, Gerard J. Criner, on behalf of the Temple University COVID-19 Research Group

## Abstract

**Background:** Surges in COVID-19 disease cases can rapidly overwhelm healthcare resources; triaging to appropriate levels of care can assist in resource planning. At the beginning of the pandemic, we developed a simple triage tool, the Temple COVID-19 Pneumonia Triage Tool (TemCOV) based on a combination of clinical and radiographic features that are readily available on presentation to categorize and predict illness severity.

**Methods:** We prospectively examined 579 sequential cases admitted to Temple University Hospital who were assigned severity categories on admission. Our primary outcome was to compare the performance of TemCOV in predicting patients who have the highest likely of admission to the ICU at 24 and at 72 hours to other standard triage tools: the National Early Warning System (NEWS), the Modified Early Warning System (MEWS) and the CURB65 score. Additional endpoints included need for invasive mechanical ventilation (IMV) within 72 hours, total hospital admission charges, and mortality.

**Results:** 26% of patients fell within our highest risk Category 4 and were more likely to require ICU admission at 24 hours (OR 11.51) and 72 hours (OR 8.6). Additionally they had the highest likelihood of needing IMV (OR 29.47) and in-hospital mortality (OR 2.37)., TemCOV performed similar to MEWS in predicting ICU admission at 24 hours (receive operator characteristic (ROC) curve area under the curve (AUC) 0.77 vs. 0.74, p=0.21) but better than NEWS2 and CURB65 (ROC AUC 0.77 vs. 0.69 and 0.77 vs. 0.64, respectively, p<0.01). While all severity scores had a weak correlation to hospital charges, the TemCOV performed the best among all severity scores measured (r=0.18); median hospital charges for Category 4 patients was $170,468 ($96,972-$487,556).

**Conclusion:** TemCOV is a simple triage score that can be used upon hospitalization in patients with COVID-19 that predicts the need for hospital resources such as ICU bed capacity, invasive mechanical ventilation and personnel staffing.

## Introduction

The coronavirus disease 2019 (COVID-19) pandemic strains healthcare resources and requires measures to improve resource allocation. With the high virulence of SARS-CoV-2 and rapid surge of COVID-19, healthcare resources became easily overwhelmed necessitating transformation of existing personnel, resources and usable spaces into surge areas.^1^ Capacity strain on a health system can lower the chances of ICU admission for both sepsis and respiratory failure independent of disease severity^2^; therefore there is an urgent need to identify methods to track ventilator and advanced respiratory resource utilization in high COVID-19 surges times through a better triage system that can be applied at patient presentation. An example of this is the COVID-GRAM risk calculator developed in China through a nation-wide analysis of hospitalized patients with COVID-19 through logistic regression.^3^ This calculator uses 10 characteristics of patients with COVID-19 to predict a composite endpoint of critical illness (ICU admission, mechanical ventilation, or death).^3^ Limitations of this scoring system include its derivation and validation in a strictly Chinese patient population and reliance on retrospective chart review to identify predictors that are statistically important but might not be clinically meaningful.

Given that few COVID-19 specific triaging scores have been developed, other traditional scoring systems have been suggested to be used to help predict hospitalization or clinical worsening in COVID-19 infection. The CURB65 was derived from a modified British Thoracic Society severity score initially as a means to identify mortality risk but later validated as a triage assessment tool. A CURB65 ≥3 has a 75% sensitivity and specificity for 30-day mortality, with high rate of mortality (22%).^4^ The National Early Warning System by the Royal College of Physicians (NEWS2) can be used to monitor the appropriateness of triage decisions in the emergency room, with ≥7 typically necessitating ICU transfer.^5^ The Modified Early Warning System (MEWS) uses vital sign and mental status assessment to predict those at risk for deterioration, with scores ≥5 associated with a 5.4-fold and 10.9-fold increased odds of death and ICU admission, respectively.^6^ The Pneumonia Severity Index (PSI) can predict those at low risk who could be managed as outpatients at severity class ≤III.^7^ However, none of these scoring systems incorporate the need for ventilatory or hemodynamic support devices, personnel staffing or types of patient care locations that are important to monitor in pandemic situations where patient surge can create high demand for similar limited resources. Adequate supplies of needed equipment, personnel and space for patient care are needed to be monitored real-time to ensure that the hospital supply chain command can project the clinical needs of a critically ill patient population to ensure an adequate supply of vital resources.

The COVID-19 pandemic offered a unique opportunity at Temple University Hospital to implement a triage system for assessing critical care needs during surges of hospitalized patients to plan rapid resource allocation. Our triage score the Temple COVID-19 Pneumonia Triage Tool (TemCOV) was developed to incorporate vital signs, type and severity of patient symptoms, the need for respiratory and hemodynamic equipment to anticipate personnel needs and necessity to convert general hospital space for critical care capability. TemCOV was developed with the primary intent to utilize data that can be ascertained immediately at patient presentation. It was conceived in mid-February 2020, before the initial surge of cases in the Philadelphia area. We present here a prospective evaluation of TemCOV in triaging patients with suspected COVID-19 infection upon presentation who required intensive care management or early consideration for COVID-19-specific therapies.

## Methods

### Patient selection

We reviewed 647 consecutive patients suspected with COVID-19 infection who were admitted to Temple University Hospital (Philadelphia, PA), a tertiary care hospital. Patients were admitted between March 14^th^ 2020 to June 3^rd^ 2020, with the majority presenting in March and April (93.7%). Data was manually extracted when needed from our electronic medical record (Epic Systems Corporation). To be eligible, patients had to test positive for COVID-19 by RT-PCR nasal swab or be admitted for symptoms consistent with COVID-19 infection and be high suspicion based on clinical presentation (chest CT imaging plus an inflammatory biomarker profile consistent with COVID-19 infection). Clinical suspicion was used an inclusion criterion given the lack of readily available COVID-19 RT-PCR testing, delayed turnaround, and suboptimal sensitivity of the RT-PCR swab in March 2020. Eight charts were duplicates, 56 patients were excluded because the final diagnosis was secondary to another process (community acquired pneumonia, heart failure exacerbation, etc.) and an additional 4 patients were excluded because they had a prolonged hospital course for another indication before testing positive for COVID-19 infection (i.e. major surgery, lung transplantation). Waiver of informed consent was granted (Temple University IRB Protocol #27051).

### Temple COVID-19 Pneumonia Triage Tool (TemCOV)

TemCOV assesses 3 “domains” of the clinical presentation – the nature and severity of symptoms, pattern and extent of pulmonary infiltrates on admission chest CT imaging, and the type and extent of respiratory or external hemodynamic support (Table 1). Symptoms were those present on admission, while vital signs were calculated using the “worst values” within the first 6 hours of presentation; initially 24 hour values were constructed but an interim analysis of 347 showed that only 14 (4%) of patients had scores escalated during this time; therefore the time period was kept on the shorter range to allow for validation in a practical triage period.

**Table 1:** Temple COVID-19 Pneumonia Triage Tool. Final severity category is assigned by the highest score among the 3 domains based on presentation within the first 6 hours. For example: A patient presenting with a respiratory rate of 26/min (score 3) requiring nasal cannula at 6lpm (score 2) and dense consolidations on CT (score 4) would have a final severity category of 4.

| Score | Symptoms/Vitals | CT Appearance | Respiratory Support |
| --- | --- | --- | --- |
| 0 | Asymptomatic carrier | Normal | Room Air |
| 1 | Afebrile, fatigue, URI or GI symptoms only | Normal | Nasal cannula $\leq 3$lpm |
| 2 | Respiratory symptoms, RR $\leq 25$/min, HR $\leq 110$bpm, Temp $\leq 101^\circ\text{F}$ | Peripheral ground glass opacities $< 3$ lobes or single lobar consolidation | Nasal cannula 4-15lpm |
| 3 | Dyspnea, Temp $> 101^\circ\text{F}$, RR $> 25$/min, HR $> 110$bpm | Peripheral and central ground glass opacities involving $\geq 3$ lobes | High flow nasal cannula or NIV |
| 4 | Hemodynamic compromised: SBP $< 90$mmHg or requiring pressors | Diffuse ground glass opacities with consolidative changes or "crazy-paving" pattern | Invasive mechanical ventilation |

Abbreviations: bpm: beats per minutes, HR: heart rate, lpm: liters per minute, NIV: non-invasive ventilation, RR: respiratory rate, SBP: systolic blood pressure

Chest computerized tomography (CT) images were stratified to assess the extent and pattern of CT abnormalities based on a pre-planned analysis plan conceived by our thoracic radiologists: grade 1 had no pulmonary involvement, grade 2 had peripheral ground glass opacities or peripheral consolidation isolated to 1-2 lobes; grade 3 CT scans had ≥3 lobar ground glass involvement with central extension; grade 4 had diffuse infiltrates, both central and peripheral, with also suggestion of advanced lesions (i.e. crazy-paving pattern, dense consolidation). Our CT scoring system predated the categories proposed jointly by the Society of Thoracic Radiology, the American College of Radiology, and Radiological Society of North America that describes typical chest CT findings for COVID-19 related disease but not the severity of infiltrates.^8^

The patient was assigned a severity category on a 0-4 scale based on the highest score among the 3 domains. During planning for anticipated triage needs, all admitted patients had CT evaluation of disease burden. Patients in the 0-1 category were usually recommended for discharge home in the absence of extenuating or psychosocial circumstances, category 2 were usually stable for the general medial ward, categories 3 and 4 were triaged to either a step down unit or ICU setting but could be triaged to lower acuity at the discretion of the admitting team.

### Endpoints

The primary endpoint was the ability of TemCOV to predict ICU admission by 24 and 72 hours using a score of 4 as a cutoff and compared it to the other scoring systems using previously defined cutoffs for high mortality risk and/or need for ICU evaluation (7 for NEWS2^5^, 5 for MEWS^6^, and 4 for CURB65^4^). Because of low percentage patients with a CURB65 score of 4, the endpoint was instead assessed for scores ≥3. A NEWS2 cutoff of 7 has a much better sensitivity for predicting ICU admissions than the lower cutoff of 5 that typically triggers a rapid response assessment (98% vs. 69%).^9, 10^ COVID-GRAM scores were calculated for patients when all variables were present, and patients were divided into the medium and high-risk groups based on the cutoff of 138.5 as determined by the prediction model.^3^

These endpoints were chosen because of the need to quickly evaluate resource allocation in a pandemic setting. Our institution was not limited by ICU capabilities as we were able to convert a separate hospital building to provide dedicated COVID-19 ICUs, step-down units and general floor bed areas, with the capabilities to provide advanced oxygenation and hemodynamic support. Space limitations did not affect influence the decision to upgrade patients to a critical level of care. We additionally looked at the predictive value of each scoring system for need of invasive mechanical ventilation (IMV) by 24 hours, in-hospital mortality, total hospital admission charges, and number of patients requiring escalation in level of care by 72 hours. Supplementary Figure 1 displays graphically the timing of data collection, calculation of individual severity scores, and endpoints.

**Figure 1:**
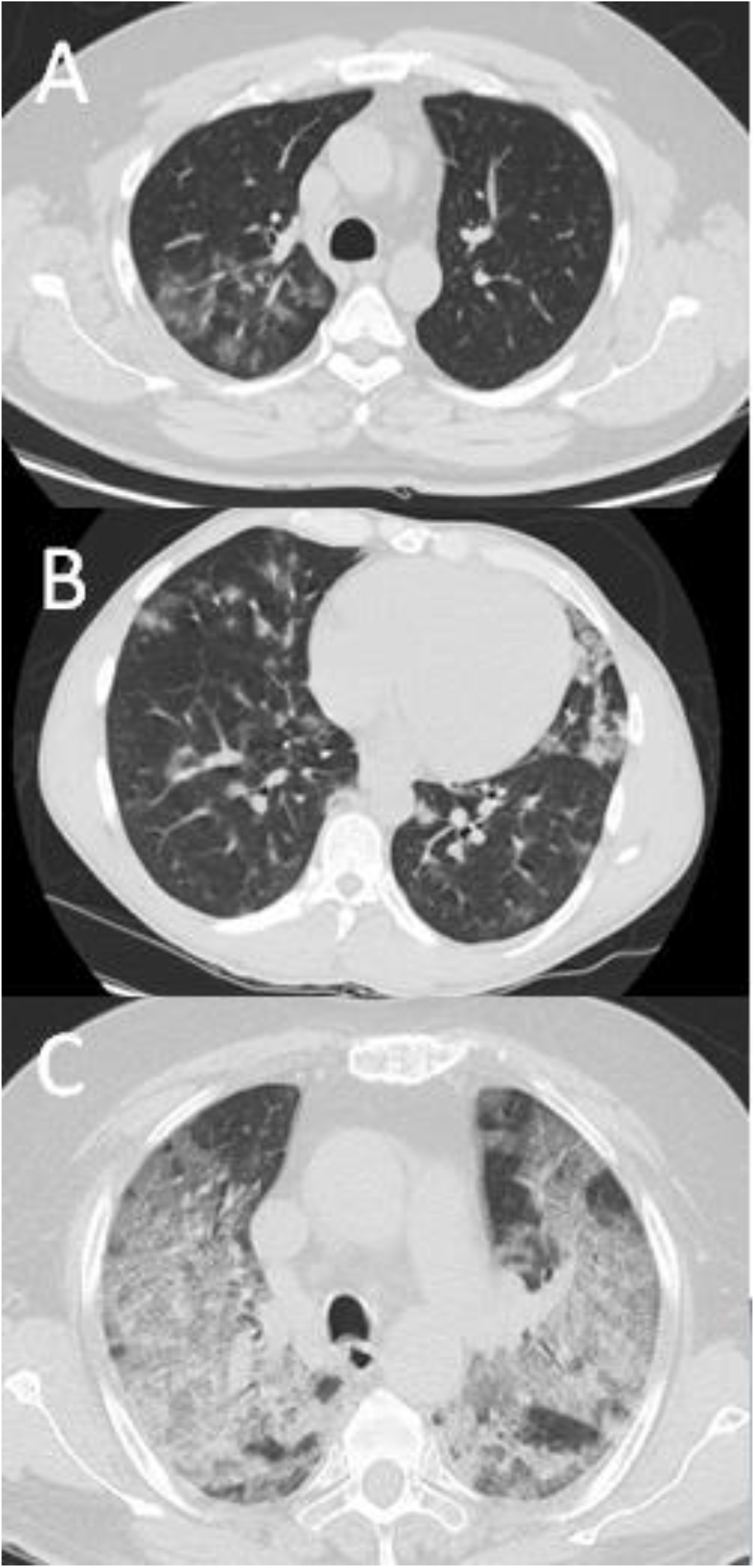
Representative CT Scans for Stages 2-4. Image A shows a Grade 2 CT: ground glass opacities isolated to the right upper lobe, all other lobes were spared. Image B shows a Grade 3 CT: central and peripheral ground glass opacities involving the right lower lobe, lingula, and left lower lobe in this representative image. Image C shows a Grade 4 CT: bilateral ground glass opacities evolving into a crazy paving pattern.

### Statistical Analysis

Descriptive statistics are reported for the entire cohort. Additionally characteristics between survivors and non-survivors, critically ill vs. those managed on the medicine floor, and those mechanically ventilated vs. those managed with other forms of oxygenation support were compared using Student’s t-test, Chi-Square, and Fisher Exact Test, where appropriate. The predictive value for each scoring system was compared using odds ratios (OR) and logistic regression at each of their respective cutoffs. Receiver-operator characteristic (ROC) curves for the primary endpoint were constructed for each scoring system and compared to the TemCOV. Hospital admission charges (in US Dollars, with interquartile range (IQR)) were compared between each severity category using Kruskal-Wallis Test for non-parametric data; for comparison between scoring systems, we converted total charges to log scale and compared Pearson’s coefficients.

The PSI and COVID-GRAM scores were assessed for patients when complete clinical data was available; therefore many patients were excluded and statistical comparisons were not performed given the reduction in power.

For all comparisons, a p-value of <0.05 was considered significant. Statistical analyses were performed with the use of Stata 14.0 (StataCorp LP, College Station, TX).

## Results

### Demographics

A total of 579 patients were included in the primary analysis; 68% percent were positive by RT-PCR. The majority (64.8%) fell into category 3 followed by category 4 (26.1%). Patients had an average age of 58.8±15.1 years, average BMI of 31.3±8kg/m^2^, and 42% were female. 89.1% had at least 1 comorbidity, the most frequently reported was hypertension (71.2%), followed by diabetes (44.7%) and psychiatric illness (21.5%); 47% were active or former smokers. 13.3% required some form of advanced respiratory therapy on admission (high flow nasal therapy (HFNT), non-invasive ventilation (NIV), or IMV). Vital sign trend within the first 24 hours and admission lab results are displayed in Table 2. Overall mortality was 12.6%.

**Table 2:** Demographic, Physiologic, and Lab Values at Presentation. Data collected over the first 24 hours of presentation.

| Variable | Total<br>(n=579) | Living<br>(n=506) | Deceased<br>(n=73) | p-value |
| --- | --- | --- | --- | --- |
| **Demographics** |  |  |  |  |
| Age, years (SD) | 58.75 (15.1) | 57.4 (15) | 68.42 (12.3) | **<0.0001** |
| Female sex, N (%) | 243 (42) | 211 (41.7) | 32 (43.8) | 0.73 |
| BMI, kg/m<sup>2</sup> (SD) | 31.33 (8) | 31.49 (8) | 30.22 (7.9) | 0.21 |
| African American, N (%) | 333 (57.6) | 289 (57.2) | 44 (60.3) | 0.06 |
| **Comorbidities** |  |  |  |  |
| Hypertension, N (%) | 412 (71.2) | 353 (69.8) | 59 (80.8) | 0.051 |
| Diabetes, N (%) | 259 (44.7) | 220 (43.5) | 39 (53.4) | 0.11 |
| Chronic Kidney Disease, N (%) | 111 (19.2) | 84 (16.6) | 27 (37) | **<0.0001** |
| Dialysis Dependent, N (%) | 43 (7.4) | 33 (6.5) | 10 (13.7) | **0.029** |
| COPD, N (%) | 71 (12.3) | 52 (10.3) | 19 (26) | **0.0001** |
| Psychiatric Comorbidities, N (%) | 124 (21.5) | 107 (21.2) | 17 (23.3) | 0.68 |
| Coronary and Peripheral Atherosclerosis, N (%) | 80 (13.8) | 63 (12.5) | 17 (23.3) | **0.012** |
| Congestive Heart Failure, N (%) | 94 (16.2) | 74 (14.6) | 20 (27.4) | **0.006** |
| Ischemic Stroke, N (%) | 69 (11.9) | 52 (10.3) | 17 (23.3) | **0.001** |
| Malignancy, N (%) | 39 (6.7) | 28 (5.5) | 11 (15.1) | **0.002** |
| **Clinical and Radiographic Features** |  |  |  |  |
| Dyspnea, N (%) | 359 (62) | 309 (61.1) | 50 (68.5) | 0.22 |
| Altered mental status, N (%) | 42 (7.3) | 26 (5.1) | 16 (21.9) | **<0.0001** |
| Systolic Blood Pressure, mmHg (SD) | 132.59 (26.6) | 133.14 (26) | 128.71 (30) | 0.18 |
| Respiratory Rate, per minute (SD) | 21.69 (5.3) | 21.42 (5.2) | 23.58 (5.6) | **0.001** |
| Heart Rate, bpm (SD) | 97.13 (19.1) | 97.02 (18.6) | 97.92 (22.3) | 0.74 |
| FiO<sub>2</sub>, % (IQR) | 21 (21-33) | 21 (21-30) | 36 (21-100) | **<0.0001** |
| O<sub>2</sub> saturation, % (SD) | 94.99 (3.6) | 95.11 (3.5) | 94.2 (4.3) | 0.078 |
| SF Ratio, % (SD) | 362.48 (122.6) | 377.15 (112.1) | 260.8 (143.5) | **<0.0001** |
| CT Grading (SD)<sup>‡</sup> | 3.03 (0.7) | 3.03 (0.7) | 3.04 (0.9) | 0.87 |
| Length of stay, days (SD) | 9.26 (9.9) | 8.32 (9.2) | 15.76 (12.4) | **<0.0001** |
| **Laboratory Values** |  |  |  |  |
| Sodium, mEq/L (SD) | 135.16 (7.6) | 134.97 (7.6) | 136.48 (7.2) | 0.11 |
| BUN, mg/dL (SD) | 24.82 (23.4) | 22.93 (21.6) | 37.92 (30.9) | **0.0001** |
| Creatinine, mg/dL (SD) | 2.14 (3.2) | 1.92 (2.8) | 3.67 (4.8) | **0.003** |
| Total Bilirubin, mg/dL (SD) | 0.7 (0.7) | 0.67 (0.5) | 0.9 (1.15) | 0.10 |
| White Blood Count, 10<sup>9</sup> cells/L (SD) | 7.41 (4.2) | 7.03 (3.8) | 10.09 (5.7) | **<0.0001** |
| Absolute Lymphocyte Count, 10<sup>3</sup> cells/μL (SD) | 1.21 (1.66) | 1.26 (1.76) | 0.93 (0.6) | **0.006** |
| Platelets, 10<sup>9</sup> cells/L (SD) | 216.69 (86.4) | 217.63 (85.1) | 210.21 (94.8) | 0.49 |
| Ferritin, μg/L (IQR) | 391.5 (184-917) | 206.5 (161-260) | 544 (260-1710) | 0.054 |
| C-Reactive Protein, mg/L (SD) | 8.15 (7) | 7.67 (6.5) | 11.39 (8.8) | **0.003** |
| Lactate Dehydrogenase, U/L (SD) | 345.21 (226) | 326.91 (192.9) | 472.98 (362.6) | **0.007** |
| D dimer, ng/mL (IQR) | 906 (501-1567) | 856 (489-1369) | 1394 (745-3296) | **0.047** |
| Fibrinogen, mg/dL (SD) | 487.04 (162.6) | 492.91 (161.2) | 450.5 (168.2) | 0.078 |

<sup>‡</sup>Semi-quantitative grading of CT burden as described in Table 1 (with representative images in Figure 1).
Abbreviations: BMI: Body Mass Index, COPD: Chronic Obstructive Pulmonary Disease, IQR: Interquartile Range, SD: Standard deviation, SF Ratio: Saturation-FiO<sub>2</sub> Ratio

### TemCOV Triage Tool

The distribution of the TemCOV categories were as follows: 11 patients (1.9%) were categories 0-1, 42 patients (7.2%) were category 2, 375 patients (64.8%) were category 3, and 151 patients (26.1%) were category 4. 127 patients in category 4 (84.1%) had a CT severity score of grade 4; additionally 17 patients of patients in this category (11.3%) required IMV and 4 patients (2.6%) required vasopressors. Distributions of physical and radiographic findings within each category are displayed in Supplementary Figure 2. Supplementary Table 1 has a further description of patients at the lowest risk category requiring admission; while all were PCR positive, none required admission for pneumonia per se, all were admitted to the regular floor, and all were discharged.

Compared to lower severity categories, category 4 was significantly associated with need for ICU admission both at 24 hours (72.5% vs. 18.6%, OR 11.51, 95% confidence interval (CI) 10.97-12.05, p<0.0001) and at 72 hours (63.5% vs. 16.8%, OR 8.6, 95% CI 8.15-9.05, p<0.0001). Likewise need for IMV at 24 hours (89.7% vs. 22.7%, OR 29.47, 95% CI 28.25-30.68, p<0.0001) and in-hospital mortality (42.5% vs. 23.7%, OR 2.37, 95% CI 1.87-2.88, p=0.0006) were more likely in the risk category 4.

In addition to predicting overall hospital outcomes, TemCOV also stratified hospital admission charges. Category 4 scores were associated with median hospital charges of $170,468 (IQR $96,972-$487,556); this was $109,400 above those incurred by Category 2 patients and $66,000 above those with Category 3 scores (Kruskal-Wallis p<0.0001). Maximum hospital charges exceeded $4 million in Category 4 patients as $3.6 million for a Category 3 patient. Box-and-whisker plots for total admission hospital charges are presented in Figure 2A.

**Figure 2:**
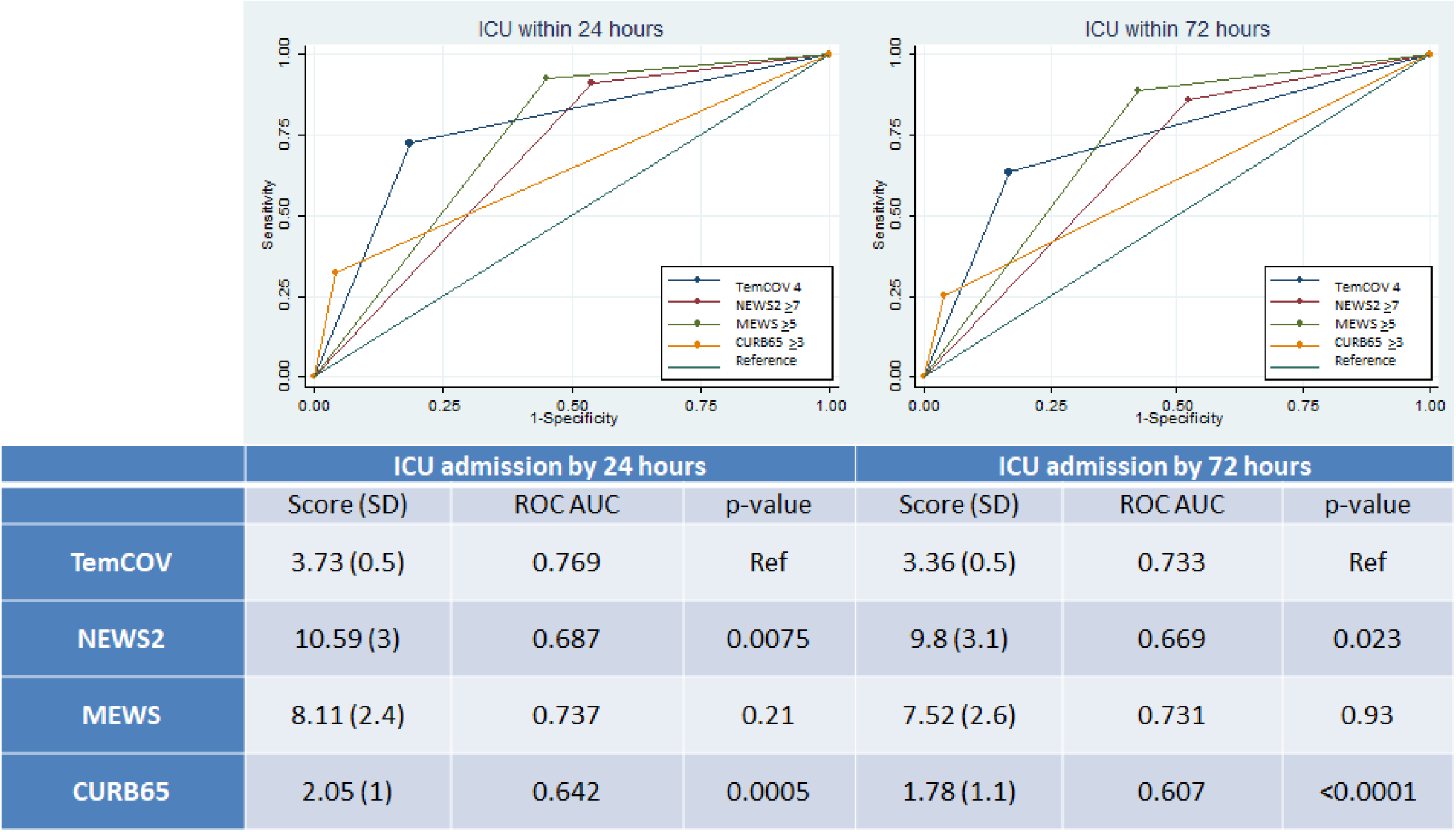
Clinical Severity Scores based on 24-hour and 72-hour Disposition. The AUC for each ROC curve shows the discriminate function for each score to predict ICU admission at specific cutoffs (TemCOV=4, NEWS2 ≥7, MEWS ≥5, and CURB65 ≥3). Bottom table displays mean scores (with standard deviations) for each severity score at 24- and 72-hours. Rationale for cutoff values is provided in Methods. Abbreviations: MEWS: Modified Early Warning Score, NEWS: National Early Warning Score, PSI: Pneumonia Severity Index, ROC AUC: Receiver Operator Characteristic Curve Area under the Curve, SD: Standard deviation, TemCOV: Temple COVID-19 Pneumonia Triage Tool

### Comparison to other severity scoring systems

Similar to our system, NEWS2≥7, MEWS≥5, and CURBS-65≥3 were significantly associated with ICU admission by 24 hours (OR 8.92, 95% CI 8.12-9.71, OR 15.02, 95% CI 14.17-15.87, OR 10.96, 95% CI 10.32-11.6, respectively, p<0.05 for all comparisons) and 72 hours (OR 5.63, 95% CI 5.07-6.19, OR 10.63, 95% CI 10.03-11.24, and OR 8.36, 95% CI 7.72-8.99, respectively, p<0.05 for all comparisons). For predicting ICU admission at 24 hours, TemCOV performed better than both the NEWS2 and CURB65 (ROC area under the curve (AUC) 0.77 vs. 0.69 and 0.77 vs. 0.64, respectively, p<0.01) and performed equally as well as the MEWS (AUC 0.77 vs. 0.74, p=0.21). Similar results were found for the predicting ICU admission by 72 hours. ROC curves with AUCs are displayed in Figure 2. Additionally, the TemCOV was better than MEWS, NEWS2, and CURB65 at predicting need for IMV at 24 hours (Figure 3). Numerically, higher risk patients using TemCOV were more likely to be transferred to a higher level of care within 72 hours after initial triage to a general medicine service compared to other those deemed higher risk based on the other triage risk calculators; however none of these comparisons reached statistical significance (Table 3).

**Table 3:** Escalation in Care Rates Between Different Scoring Systems. Number (with percentage) of patients who were upgraded to ICU after initial triage to a general medicine floor based on estimated risk category (high versus low risk). Statistical comparison for each group is in comparison to TermCOV by Fisher Exact Test.

|  |  | TemCOV | NEWS | MEWS | CURB65 |
| --- | --- | --- | --- | --- | --- |
| Low Risk | Upgrade in Care (N, %) | 20 (4.67) | 9 (3.8) | 7 (2.5) | 32 (6.02) |
|  | Fisher | Ref | 0.695 | 0.166 | 0.474 |
| High Risk | Upgrade in Care (N, %) | 15 (9.93) | 26 (7.6) | 28 (9.36) | 3 (6.38) |
|  | Fisher | Ref | 0.483 | 0.867 | 0.77 |

Abbreviations: MEWS: Modified Early Warning Score, NEWS: National Early Warning Score, TemCOV: Temple COVID-19 Pneumonia Triage Tool

**Figure 3:**
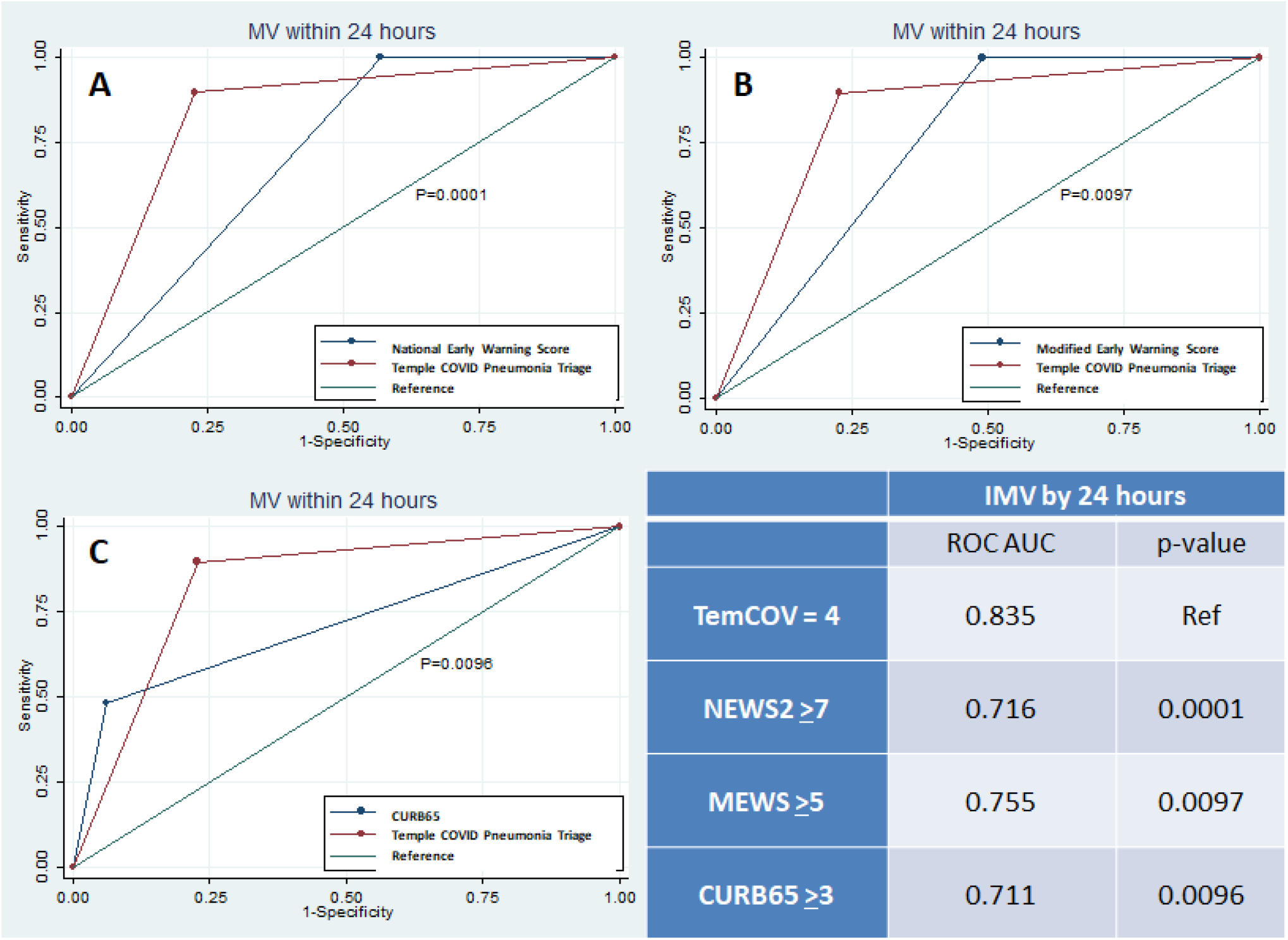
Predicting 24-hour Invasive Mechanical Ventilation by Clinical Severity Scores. Graphs display ROC curves for ability of each triage tool to predict 24 hour need for IMV for respiratory failure due to COVID-19; red ROC curve is the Temple COVID-19 Pneumonia Triage Tool in comparison to curves for NEWS2 (A), MEWS (B), and CURB65 (C). Table displays AUC for each severity score ROC curve. p-values displayed are in comparison to the TemCOV score. Rationale for cutoff values is provided in Methods. Abbreviations: IMV: Invasive Mechanical Ventilation, MEWS: Modified Early Warning Score, NEWS: National Early Warning Score, PSI: Pneumonia Severity Index, ROC AUC: Receiver Operator Characteristic Curve Area under the Curve, TemCOV: Temple COVID-19 Pneumonia Triage Tool

All severity scores had low but positive correlation between increasing severity measures and the log transformation of hospital admission charges. Of these, the TemCOV score had the highest numerically (r=0.18) and statistically greater than NEWS (r=0.032, p=0.002) but similar to the correlations for MEWS and CURB65 (r=0.17 and r=0.15, p>0.05 for both). Box and whisker plots for hospital charges among the different severity scores are presented in Figure 4.

**Figure 4:**
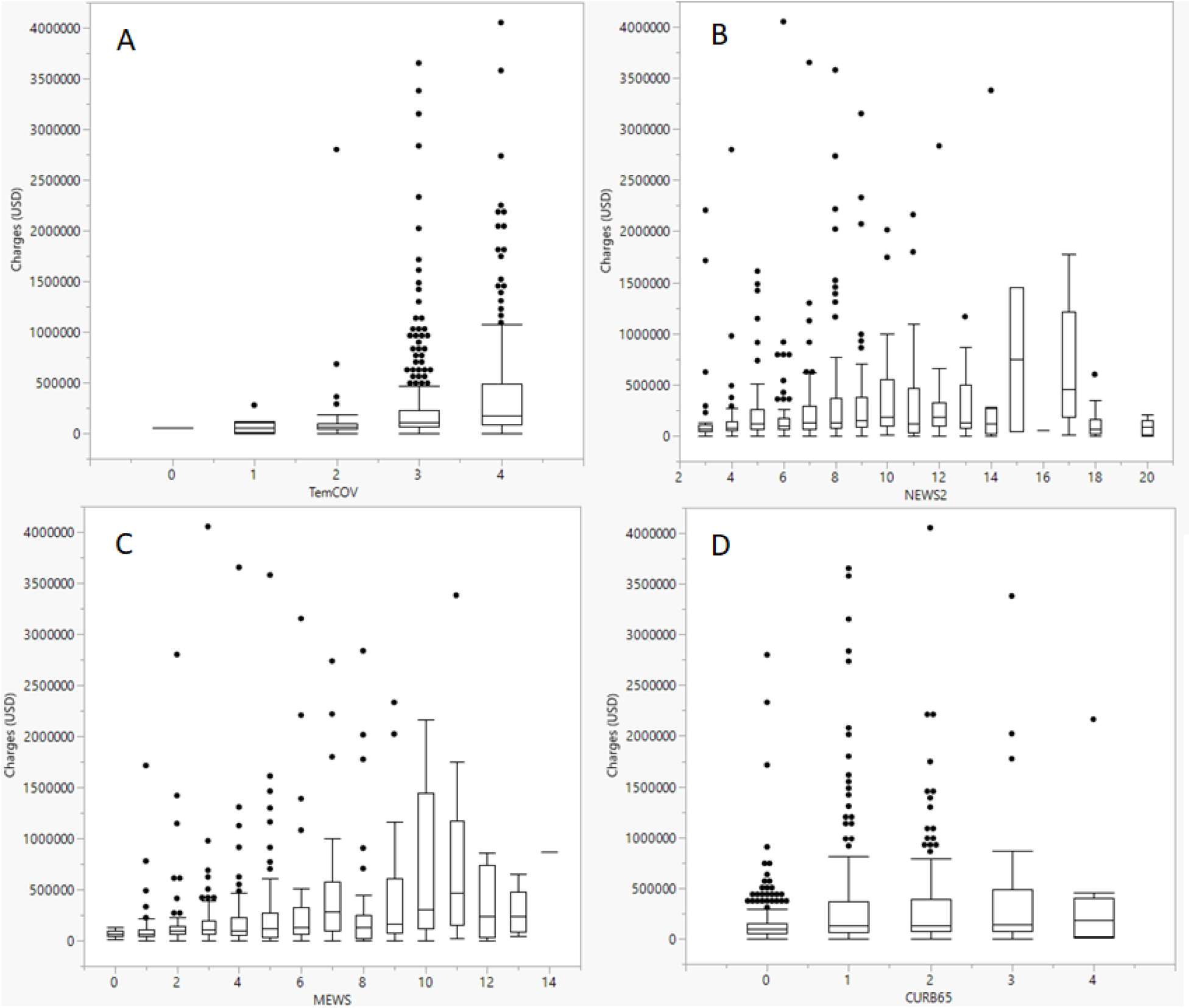
Distribution of Hospital Charges by Severity Score. Displayed are box-and-whisker plots for corresponding total admission hospital charges for each severity scale (A:TemCOV, B:NEWS2, C:MEWS, D:CURB65) by scoring. Charges are presented in USD amount. Abbreviations: MEWS: Modified Early Warning Score, NEWS: National Early Warning Score, ROC AUC: Receiver Operator Characteristic Curve Area under the Curve, SD: Standard deviation, TemCOV: Temple COVID-19 Pneumonia Triage Tool, USD: US Dollar

### COVID-GRAM and PSI

Because of unavailability of arterial blood gases or direct bilirubin, the PSI and COVID-GRAM scores were only available in 57 (9.8%) and 87 (15%) patients, respectively, so could not be compared to the TemCOV and other triage tools; only exploratory results are presented here. Thirty-nine patients had a PSI score ≥4 (average score 118.12±39.7) with a mortality of 43.6% compared to 16.7% those with scores <4 (p=0.073). Average COVID-GRAM score was 121.2±30.8 with 28.7% falling into the high risk category; mortality was significantly higher in this group (64.3% vs. 21.9%, p=0.006). Need for ICU at 72 hours was significantly higher in patients with higher severity scores (87.1% vs. 61.1%, p=0.037 for PSI, and 44% vs. 19.4%, p=0.03 for COVID-GRAM).

## Discussion

We have shown that a simple scoring system TemCOV can quickly and easily predict patients who would require higher level of care immediately upon admission for COVID-19 pneumonia. TemCOV is independent of comorbidities despite nearly 90% of our population having at least one baseline medical condition. TemCOV’s ability to distinguish risk for in-hospital mortality was modest; our overall mortality was 12%, lower than initial reports from the COVID-19 pandemic^11, 12^ and more in-line with recent estimates.^3, 13^ Our score uses some clinical vital sign parameters that are incorporated within other scoring systems, such as the COVID-19 Severity Index, that use respiratory rate, pulse oximetry, and O2 flow rate to evaluate for 24-hour respiratory decompensation.^14^ In contrast to this score, TemCOV included mechanical ventilation and pressor use to provide clear guidance for triage, plan staff coverage, and track resource utilization. However, the number of category 4 patients who required either of these interventions was low (11% and 3%, respectively); instead many more fell into the highest risk category based on CT appearance and our findings underscore the importance of radiographic disease burden and its value in prediction of hypoxemic respiratory failure. Prediction scores based on CT disease burden can discriminate between those that present with mild disease compared to those who could develop critical illness.^15^ Semi-quantitative CT involvement and not individual CT features (such as consolidation or air bronchograms) have been selected through machine learning for use in predictive modeling, but CT appearance was ultimately not incorporated in Wu and colleague’s risk calculator.^16^ Category 4 severity was associated with an 11-fold increased risk for ICU transfer within 24 hours of admission and an 8-fold increased risk by 72 hours. Not only does this translate to patient outcomes but hospital resource utilization as demonstrated by the increase in hospital charges, a surrogate for staffing and equipment needs.

Traditional scores of severity of illness can predict in-hospital mortality for COVID-19 patients^10, 12, 13, 17, 18^ although evidence is limited to single-center experience with small sample sizes and fewer provide validation in a US center. A previous study of 105 patients in China comparing MEWS with the Rapid Emergency Medicine Score, the optimal cutoff for MEWS was found to be 2 for mortality, albeit with the modest sensitivity and specificity (68% and 65%, respectively);^19^ our present study found a higher mean MEWS score among survivors (4.6). Adding an age cutoff ≥65 years in this previous study only slightly improved its predictive value.^19^ This discrepancy may be related to difference in time course of our data collection (we examined the first 24 hours of admission for MEWS calculation) and a higher comorbidity burden overall. The CURB65 has also been studied for its role in the care of patients with COVID-19 pneumonia. A single-center French study showed HR 4.18 of scores 3-5 in meeting the composite outcome (ICU need, high flow oxygen, noninvasive or invasive ventilation, death), but still 20% of patients with a CURB65 <2 met the primary endpoint including 9% mortality at 14 days.^20^ A Turkish study comparing PSI to CURB65 showed superiority of the former in predicting 30-day mortality, but still excellent predictive value between both models (AUC for ROC 0.88 vs. 0.9).^13^ The mortality with a CURB65 ≥2 was 30.5% compared to 2.7% with lower scores; likewise higher PSI risk categories had mortality upwards of 28-65%.^13^ A more recent comparative study evaluating most of the scoring systems including in our current study confirmed excellent predictive value for in-hospital mortality with AUC for all exceeding 0.7. CURB-65, PSI, and NEWS2 demonstrated excellent (>90%) negative predictive value but low-moderate positive predictive value for predicting death at cutoffs of 2, 3, and 5, respectively. ^18^

The original validation cohort of the COVID-GRAM score in China^3^ and more recent risk calculator developed by Wu *et al*. from Chinese and European populations^16^ had lower morbidity burden compared to ours and other^11^ US populations. Clinical scoring systems in general tend to show diminished predictive value outside of their derivation population.^21^ Although the COVID-GRAM maintained performance in predicting death in our cohort of COVID-19 patients, the inclusion of factors not routinely checked excluded application in nearly 85% of our cohort. If these laboratory factors are not readily available, the model cannot initially triage patients to appropriate levels of care. Components with low frequencies and questionable clinical significance (i.e. hemoptysis reported in 1.4% of our cohort and unconsciousness seen in only 2.9%) could lead to over-fitting of prediction models and reduce performance when more widely applied.^22, 23^ Specific COVID-19 prediction models that can be utilized at point-of-care in larger and more diverse populations are needed. Shang et al. also recently developed a statistical model for predicting death in COVID-19 that utilized procalcitonin,^12^ which may not be widely or readily available in some institutions. Death was the outcome of interest in this study and occurred about 3 weeks into their disease course;^12^ however progression to end-organ complications including respiratory failure, acute kidney injury, and sepsis developed much earlier (median 11-13 days) after symptoms onset.^12^ Therefore, we focused on short-term endpoints to validate our scoring system to make TemCOV clinically useful for triage and resource management purposes upon admission.

TemCOV scoring is simple to use and performed comparably against more cumbersome systems. While our scoring system did not include any typical inflammatory markers of COVID-19 related cytokine storm, we intended to create a triage tool that did not rely on specialized markers which may not be immediately available at all hospitals. There are conflicting results on the use of biomarkers in the current literature of COVID-19-specific triage tools with some studies failing to demonstrate improved prognostic accuracy using CRP or procalcitonin,^13, 14^ while others incorporate such markers to increase predictive value.^16^ We chose 72 hours for our primary outcome, which is within the timeframe tested by other triage protocols, because more long-term endpoints (i.e. ICU transfer by discharge, mortality) are markers of prognostication and not ICU resource needs on admission. Limitations of our study include the validation of scores in a single health care system. We also recognize that our score does not take into account baseline comorbidities which can play a major role in the clinical outcomes and has been advocated by joint task forces.^1^ Some of our patients on high flow nasal cannula therapy were managed on the general medical floor, which may be atypical for some institutions, but the incidence of this was low overall (6%).

## Conclusion

TemCOV is a simple tool for triage immediately upon hospitalization in patients with COVID-19. It can be tabulated within hours of initial presentation and updated throughout a patient’s hospital course and used to predict the need for hospitalization resources-ICU bed capacity, invasive mechanical ventilation and other forms of respiratory equipment and personnel staffing. Higher TemCOV scores should initiate prompt clinical response and consideration for more frequent monitoring. TemCOV needs to be validated by others and given its nonspecific scoring characteristics, can also be studied in other types of viral pneumonias.

## Data Availability

Data was extracted and anonymized from our electronic medical record and stored on password-protected computers at the Temple University Health System.

## Acknowledgements

The following are acknowledging for also contributing to data collection: Rohit Soans, MD, Nicole Patlakh, BSc, Ryan Townsend, MD, Tse-Shuen Ku, MD, Melinda Darnell, MD, Zachariah Dorey-Stein, MD, Catherine Myers, MD, Ibraheem Yousef, MD, and Stephanie Jeong, MD. We would also like to acknowledge Abhi Rastogi, MBA, MIS and Christopher Snyder, MBA for assistance with compiling hospital charge data.

## List of Abbreviations

CI: Confidence Interval
COVID-19: Coronavirus disease 2019
CT: Computerized Tomography
IMV: Invasive Mechanical Ventilation
IQR: Interquartile Range
MEWS: Modified Early Warning System
NEWS2: National Early Warning System 2
OR: Odds Ratio
PSI: Pneumonia Severity Index
ROC: Receiver-operator Characteristic
TemCOV: Temple COVID-19 Pneumonia Triage Tool
USD: US Dollars

