## Supplementary material for "Utility of a Clinical Scoring System for Point of Care Triaging in COVID-19 Pneumonia": Online Supplement

**Online Supplementary Information**

Andrew J Gangemi, MD^1*^, Rohit Gupta, MBBS^1*^, Gustavo Fernandez-Romero, MD^1^, Huaqing Zhao, PhD, MS^2^, Maulin Patel, MD^1^, Junad Chowdhury, MD^1^, Massa Zantah MD^1^, Matthew Zheng, MD^1^, Osheen Abramian, MD^7^, Stephen Codella, MD^1^, Linda Vien, MD^3^, Eduardo Dominguez-Castillo, MD^1^, Timothy Buckey, MD^3^, Charles Earley, MD^3^, Jourdan Frankovich, MD^3^, Mali Jurkowski, MD^3^, Zachary Jurkowski, MD^3^, Jenny O’Brien, MD, PhD^3^, Nanzhou Guo, CRNP^1^, Paige Stanley, PA-C^1^, Brenton Halsey, MD, MA^3^, Jasleen Kahlon, MD^3^, Navjot Kaur, MD^1^, Roman Prosniak, MD^1^, Maruti Kumaran, MD, MBBS^4^, Chandra Dass, MBBS^4^, David Fleece, MD^5^, Michael R Jacobs, PharmD^1,6^, and Gerard J. Criner MD^1^ on behalf of the Temple University COVID-19 Research Group

Lewis Katz School of Medicine at Temple University, Philadelphia, PA

^1^Department of Thoracic Medicine and Surgery

^2^Department of Clinical Sciences

^3^Department of Internal Medicine

^4^Department of Radiology

^5^Department of Clinical Pediatrics

^6^Department of Pharmacy Practice

Cooper University Hospital, Camden, NJ

^7^Department of Pulmonary and Interventional Pulmonary Medicine

*Contributed Equally

**Collaborators in the Temple University COVID-19 Research Group:**

Aaron Mishkin; Abbas Abbas; Abhijit S Pathak; Abhinav Rastogi; Adam Diamond; Aditi Satti; Adria Simon; Ahmed Soliman; Alan Braveman; Albert J Mamary; Aloknath Pandya; Amy Goldberg; Amy Kambo; Andrew Gangemi; Anjali Vaidya; Ann Davison; Anuj Basil; Arthur Lau; Arundathi Jayatilleke; Bakhos, Charles T; Bill Cornwell; Brent Lawrence; Brianna Sanguily; Brittany Corso; Carla Grabianowski; Carly Sedlock; Catherine Myers; Charles Bakhos; Chenna Kesava; Reddy Mandapati; Cherie Erkmen; Chethan Gangireddy; Chih-ru Lin; Christopher T Burks; Claire Raab; Crabbe, Deborah; Crystal Chen; Daniel Edmundowicz; Daniel Sacher; Daniel Salerno; Daniele Simon; David Ambrose; David Ciccolella; Debra Gillman; Dolores Fehrle; Dominic Morano; Donnalynn Bassler; Edmund Cronin; Eduardo Dominguez; Ekam Randhawa; Ekamjeet Randhawa; Eman Hamad; Eneida Male; Erin Narewski; Francis Cordova; Frederic Jaffe; Frederich Kueppers; Fusun Dikengil; Galli, Jonathan; Gangemi, Andrew; Garfield, Jamie; Gayle Jones; Gennaro Calendo; Gerard Criner; Gilbert D’Alonzo; Ginny Marmolejos; Gordon, Matthew; Gregory Millio; Gupta, Rohit; Gustavo Fernandez; Hannah Simborio; Harwood Scott; Heidi Shore-Brown; Hernan Alvarado; Ho-Man Yeung; Ibraheem Yousef; Ifeoma Oriaku; Iris Jung-won Lee; Isaac Whitman; James Brown; Jamie L. Garfield; Janpreet Mokha; Jason Gallagher; Jeffrey Stewart; Jenna Murray; Jessica Tang; Jeyssa Gonzalez; Jichuan Wu; Jiji Thomas; Jim Murrett; Joanna Beros; John M. Travaline; Jolly Varghese; Jordan Senchak; Joseph Lambert; Joseph Ramzy; Joshua Cooper;Jun Song; Junad Chowdhury; Justin Levinson; Kaitlin Kennedy; Karim B Ahmed; Karim Loukmane; Karthik Shenoy; Kathleen Brennan; Keith Johnson; Kevin Carney; Kevin Lu; Kraftin Schreyer; Kristin Criner; Kumaran, Maruti; Lauren Miller; Laurie Jameson; Laurie Johnson; Laurie Kilpatrick; Lawrence Brent; Lii-Yoong Criner; Lily Zhang; Lindsay K Mcgann; Llera A Samuels; Marc Diamond; Margaret Kerper; Maria Vega Sanchez; Mariola Marcinkienwicz; Maritza Pedlar; Mark Aksoy; Mark Weir; Marla R. Wolfson; Marla Wolfson; Marron, Robert; Martin Keane; Massa Zantah; Mathew Zheng; Matthew Delfiner; Matthew Gordon; Maulin Patel; Megan Healy; Melinda Darnell; Melissa Navaro; Meredith A. Brisco-Bacik; Michael Bromberg; Michael Gannon; Michael Jacobs; Mira Mandal; Nanzhou Gou; Narewski, Erin; Nathaniel Marchetti; Nathaniel Xander; Navjot Kaur; Neil Nadpara; Nicole Desai; Nicole Mills; Norihisa Shigemura; Ohoud Rehbini; Oisin O’Corragain; Omar Sheriff; Oneida Arosarena; Osheen Abramian; Paige Stanley; Parag Desai; Parth Rali; Patrick Mulhal; lPravin Patil; Priju Varghese; Puja Dubal; Puja Patel; Rachael Blair; Rajagopalan Rengan; Rami Alashram; Randol Hooper; Rebecca A Armbruster; Regina Sheriden; Robert Marron; Rogers Thomas; Rohit Gupta; Rohit Soans; Roman Petrov; Roman Prosniak; Romulo Fajardo; Ruchi Bhutani; Ryan Townsend; Sabrina Islam; Samantha Pettigrew; Samantha Wallace; Sameep Sehgal; Samuel Krachman; Santosh Dhungana; Sarah Hoang; Sean Duffy; Seema Ran; Sheila Weaver; Shelu Benny; Sheril George; Shuang Sun; Shubhra Srivastava-Malhotra; Stephanie Brictson; Stephanie Spivack; Stephanie Tittaferrante; Stephanie Yerkes; Stephen Priest; Steve Codella; Steven G Kelsen; Steven Houser; Steven Verga; Sudhir Bolla; Sudhir Kotnala; Sunil Karhadkar; Sylvia Johnson; Tahseen Shariff; Tammy Jacobs; Thomas Hooper; Tom Rogers; Tony S. Reed; Tse-Shuen Ku; Uma Sajjan; Victor Kim; Whitney Cabey; William Shapiro; Wissam Chatila; Wuyan Li; Zachariah Dorey-Stein; Zachary D Repanshek.

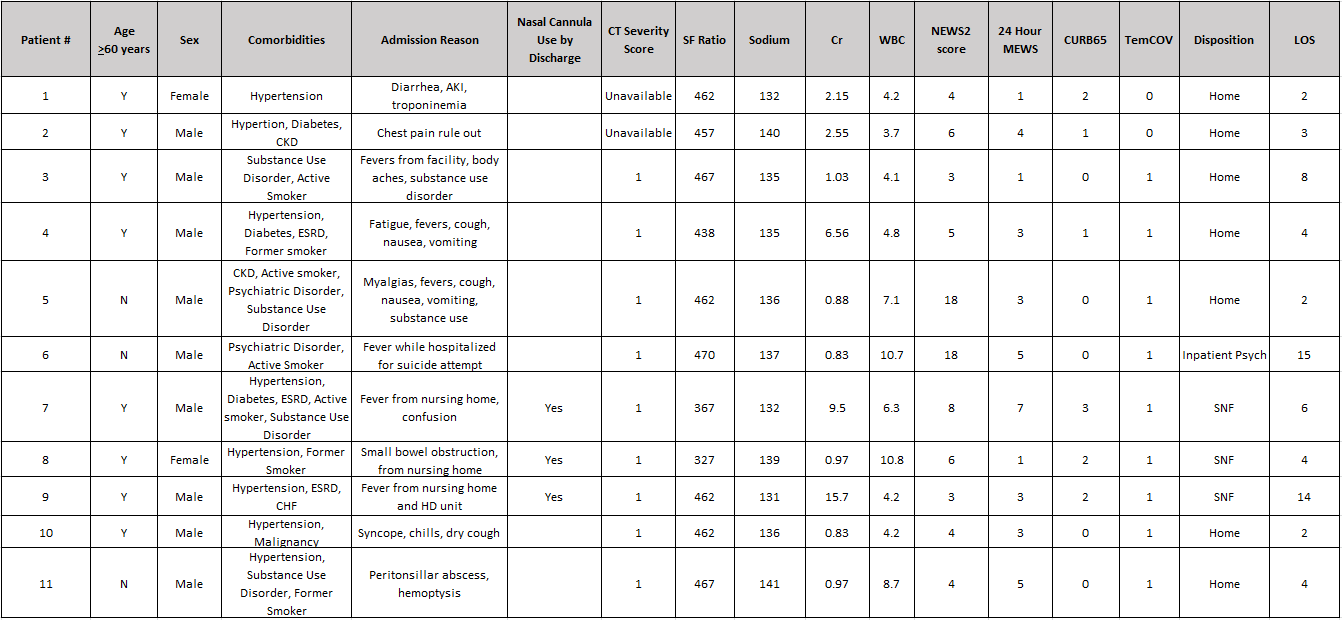

**Supplementary Table 1: Characteristics of Admitted TemCOV Scores 0-1 Patients.** All patients were discharged from the hospital and none required ICU level of care services.

Abbreviations: AKI: Acute kidney injury, CHF: Congestive heart failure, CKD: Chronic kidney disease, Cr: Creatinine, ESRD: End stage renal disease, LOS: Length of stay, SF Ratio: Saturation/FiO2 ratio, SNF: Skilled nursing facility, WBC: White blood count

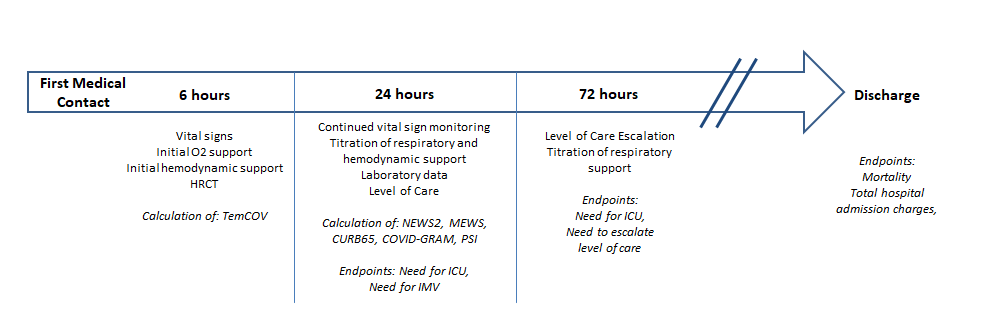

**Supplementary Figure 1: Timeline of Patient Admission Course**

Upon first medical contact and for the first 6 hours, vital sign data, initial O2 support, and HRCT data was collected allowing calculation of the TemCOV score. Over the following 24 hours, “worst” vital signs, highest hemodynamicand respiratory support, and laboratory data allowed calculation of remainder of severity scores. Timing of individual endpoints during admission and at discharge are indicated.

Abbreviations: HRCT: High resolution Computed Tomography of Chest, IMV: Invasive Mechanical Ventilation, MEWS: Modified Early Warning Score, NEWS: National Early Warning Score, PSI: Pneumonia Severity Index, TemCOV: Temple COVID-19 Pneumonia Triage Tool

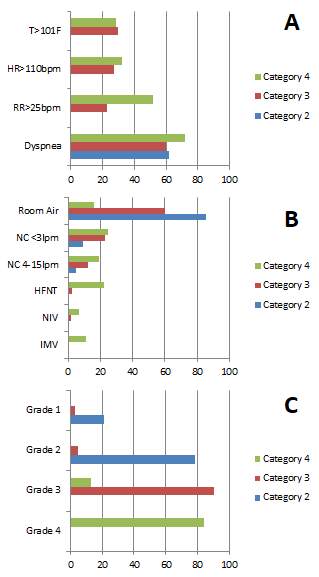

**Supplementary Figure 2: Distribution of Presenting Signs by Temple COVID-19 Pneumonia Triage Tool.** Displayed is the percent of the patients within each severity category who met physiologic cutoffs (A), required oxygenation support (B), and distribution of CT severity grades (C).

Abbreviations: HFNT: High flow nasal therapy, HR: Heart Rate, IMV: Invasive mechanical ventilation, NC: Nasal cannula, NIV: Non-invasive ventilation, RR: Respiratory Rate
